# Forecasting COVID-19 disease cases using the SARIMA-NNAR hybrid model

**DOI:** 10.1101/2021.04.26.21256108

**Authors:** İbrahim Demir, Murat Kirişci

## Abstract

**Background:** COVID-19 is a new disease that is associated with high morbidity that has spread around the world. Credible estimating is crucial for control and prevention. Nowadays, hybrid models have become popular, and these models have been widely implemented. Better estimation accuracy may be attained using time-series models. Thus, our aim is to forecast the number of COVID-19 cases with time-series models.

**Objective:** Using time-series models to predict deaths due to COVID-19.

**Design:** SARIMA, NNAR, and SARIMA-NNAR hybrid time series models were used using the COVID-19 information of the Republic of Turkey Health Ministry.

**Participants:** We analyzed data on COVID-19 in Turkey from March 11, 2020 to February 22, 2021.

**Main Measures:** Daily numbers of COVID-19 confirmed cases and deaths.

**Materials and methods:** We fitted a seasonal autoregressive integrated moving average (SARIMA)–neural network nonlinear autoregressive (NNAR) hybrid model with COVID-19 monthly cases from March 11, 2020, to February 22, 2021, in Turkey. Additionally, a SARIMA model, an NNAR model, and a SARIMA–NNAR hybrid model were established for comparison and estimation.

**Results:** The RMSE, MAE, and MAPE values of the NNAR model were obtained the lowest in the training set and the validation set. Thus, the NNAR model demonstrates excellent performance whether in fitting or forecasting compared with other models.

**Conclusions:** The NNAR model that fits this study is the most suitable for estimating the number of deaths due to COVID-19. Hence, it will facilitate the prevention and control of COVID-19.

## 1. Introduction

The latest threat to global health is the ongoing outbreak of respiratory disease that was recently given the name Coronavirus Disease 2019 (Covid-19). Covid-19 was recognized in December 2019[1]. It was rapidly shown to be caused by a novel coronavirus that is structurally related to the virus that causes severe acute respiratory syndrome (SARS). As in two preceding instances of emergence of coronavirus disease in the past 18 years[2] — SARS (2002 and 2003) and the Middle East respiratory syndrome (MERS) (2012 to the present) — the Covid-19 outbreak has posed critical challenges for the public health, research, and medical communities.

Public health groups are monitoring the pandemic and posting updates on their websites. These groups have also issued recommendations for preventing and treating the illness. As of now, researchers know that the new coronavirus is spread through droplets released into the air when an infected person coughs or sneezes. The droplets generally do not travel more than 30 centimeters, and they fall to the ground (or onto surfaces) in a few seconds.

Due to the high infectivity of the virus and the lack of immunity in the human population, the epidemic grows exponentially without intervention, and thus can greatly stress the public health system and bring enormous the disruption to economy and society. Thus, a crucial task facing every country is to reduce the transmission rate and flatten the (infection) curve. Additionally, each country is at a different stage of the epidemic and it is essential for countries to understand their own pattern of virus growth, as such information is critical for important policy decisions such as extending lockdown or reopening. To (at least partially) answer these questions, a natural step is to analyze the trajectory of the infection curve of COVID-19 since the initial outbreak in each country.

The virus can exacerbate through the respiratory tract and enter into a person’s lungs. This causes damage to the air sacs or alveoli, which can fill with fluid. This progression then constraints a person’s ability to take in oxygen. Continuous oxygen deprivation can damage many of the body’s organs, causing kidney failure, heart attacks, and other life-threatening conditions. People who have pre-existing conditions such as cancer, diabetes, high blood pressure, kidney or liver disease, including but not limited to asthma are at most risk of COVID-19 pneumonia. People over the age of 65 years are more prone to the intense effects of this disease. The disease has turned into a widespread pandemic where the cases and deaths seem to surge rapidly day by day.

Time series analysis is a scientific quantitative prediction of the future trend of diseases based on historical data and time variables. It is a quantitative analysis method that does not consider the influence of complex factors. In this study, Seasonal Auto-Regressive Moving Average (SARIMA) and Neural Network Auto-Regressive (NNAR) were used were applied on the predicting COVID-19 disease cases.

A number of authors have published papers on models for the occurrence of Coronavirus pandemic. In [10], the trajectory of the cumulative confirmed cases and deaths of COVID-19 (in log scale) via a piecewise linear trend model is given a model. Peipei and et al [11] integrated the most updated COVID-19 epidemiological data before June 16, 2020, into the Logistic model to fit the cap of epidemic trend. A machine learning-based time series prediction model to derive the epidemic curve and predict the trend of the epidemic is produced in [11]. In [12], the Bayesian structural time series (BSTS) models to investigate the temporal dynamics of COVID-19 in the top five affected countries around the world in the time window March 1, 2020, to June 29, 2020, are used. In the study of He et al.[13], an analysis was made between deaths due to COVID-19 and environmental conditions (including ambient pollutants and meteoroidal parameters). Ribeiro et al. [14] developed efficient short-term forecasting models for forecasting the number of future cases by using an autoregressive integrated moving average (ARIMA), cubist regression (CUBIST), random forest (RF), ridge regression (RIDGE), support vector regression (SVR) and stacking-ensemble learning models for evaluating in the task of time series forecasting with one, three, and six days ahead of the COVID-19 cumulative confirmed cases in ten Brazilian states with a high daily incidence. Topaç et al. [4] analyzed the cognitions, feelings, and thoughts of early childhood children who stayed at home during the quarantine process due to coronavirus with multiple correspondence analyses. In [5], the risks of the effects of quarantine status related to the COVID-19 pandemic on the cognition and behavior of children staying at home are evaluated.

The Covid-19 outbreak is a stark reminder of the ongoing challenge of emerging and reemerging infectious pathogens and the need for constant surveillance, prompt diagnosis, and robust research to understand the basic biology of new organisms and our susceptibilities to them, as well as to develop effective countermeasures.

The purpose of this study was to compare the time series models to forecast the COVID-19 incidence epidemic and it is the prediction of deaths due to Covid-19 in Turkey. For better forecasting performance, a comparison of time-series models to forecast infectious disease was studied. The results from this study will be helpful to predict future COVID-19 incidence epidemics and optimize COVID-19 control and intervention using the predictions as reference information.

## 2. Methods

### 2.1. Dataset

Data were taken in COVID-19 Information Page of Republic of Turkey Health Ministry. The dataset comprises of both confirmed and death cases only as variables refer to daily cases and covers the period from March 11, 2020, up until February 22, 2021 [7].

The R programming language [8] has been used to carry out the all analyses involved in the present investigation.

### 2.2. Time-Series Approaches

#### 2.2.1. SARIMA

Autoregressive Integrated Moving Average, or ARIMA, is one of the most widely used forecasting methods for univariate time series data forecasting. Although the method can handle data with a trend, it does not support time series with a seasonal component. An extension to ARIMA that supports the direct modeling of the seasonal component of the series is called SARIMA. In this tutorial, you will discover the Seasonal Autoregressive Integrated Moving Average, or SARIMA, method for time series forecasting with univariate data containing trends and seasonality. Seasonal Autoregressive Integrated Moving Average, SARIMA or Seasonal ARIMA, is an extension of ARIMA that explicitly supports univariate time series data with a seasonal component. It adds three new hyper parameters to specify the auto regression (AR), differencing (I) and moving average (MA) for the seasonal component of the series, as well as an additional parameter for the period of the seasonality.

For a seasonal series with s periods per year, the SARIMA(p,d,q)(P,D,Q)s models are used. Thus, having a *B*^*s*^ operator such that *B*^*s*^*X*_*t*_ = *X*_*t*−*s*_ and since the seasonal difference can be written as (*X*_*t*_ − *X*_*t*−*s*_) = (1 − *B*^*s*^)*X*_*t*_, a SARIMA model with (p,d,q) non-seasonal order terms and (P,D,Q) seasonal order terms. A time series {*Z*_*t*_. : 1,2, …, *k*} is generated by SARIMA (*p, d, q*)(*P, D, Q*)*s* process of Box and Jenkins time series model if

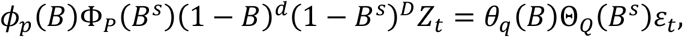

where p,d,q,P,D,Q are integers, s is the season length;

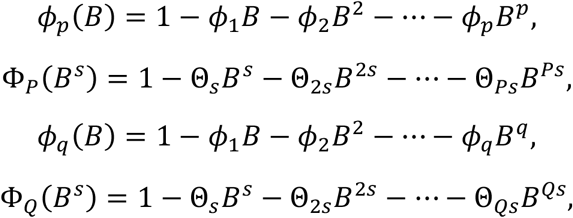

are polynomials in B, where B is the backward shift operator, *ε*_*t*_ is the estimated residual at time t, d is the number of regular differences, D is the number of seasonal differences, *Z*_*t*_ denotes the observed value at time t (t=1,2,..,k).

Fitting a SARIMA model to data involves the following four-steps iterative cycles:)a= identify the SARIMA (*p, d, q*)(*P, D, Q*)’s structure; (b) estimate unknown parameters; (c) perform goodness-fit tests on the estimated residuals; (d) forecast future outcomes based on the known data. The fitting of SARIMA models is a challenging task.

#### 2.2.2. NNAR

Neural networks are used for complex non-linear forecasting. With time series data, lagged values of the time series can be used as inputs to a neural network, just as we used lagged values in a linear autoregression model. We call this a neural network autoregression or NNAR model. NNAR is generally describes by NNAR(p,k) where p is lagged inputs and k a number of hidden layers. Also, NNAR(p,P,k) is the general denotation of seasonal NNAR. For example, a NNAR(9,5) model is a neural network with last nine observations (*y*_*t*−1_, *y*_*t*−2_, …, *y*_*t*−9_) used as inputs for forecasting the output *y*_*t*_ and with five neurons in the hidden layer. A NNAR(p,0) model is equivalent to an ARIMA(p,0,0) model, but without the restrictions on the parameters to ensure stationary. With seasonal data, it is useful to also add the last observed values from the same season as inputs. For example, an NNAR(3,1,2)_12_ model has inputs *y*_*t*−1_, *y*_*t*−2_, *y*_*t*−3_ and *y*_*t*−12_, and two neurons in the hidden layer. More generally, an NNAR(p,P,k)_m_ model has inputs (*y*_*t*−1_, *y*_*t*−2_, …, *y*_*t*−*p*_, *y*_*t*−*m*_, *y*_*t*−2*m*_, *y*_*t*−*Pm*_) and k neurons in the hidden layer. A NNAR(p,P,0)_m_ model is equivalent to an SARIMA(p,0,0)(P,0,0)_m_ model but without restrictions on the parameters that ensure stationary.

The NNAR model is a feedforward neural network which involves a linear combination function an d a activation function. The formations of these function are defined as,

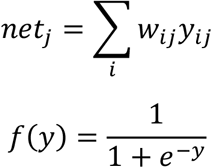

#### 2.2.3. Hybrid Model

A time series can be considered as comprising a linear autocorrelation structure and a non-linear component. The SARIMA model and the NNAR are methodologies that predict future values using historically observed data, and are suitable for linear and non-linear problems, respectively. We used the hybrid model combining the SARIMA model (linear approach) and the NNAR (non-linear approach) for this study.

In the first place, a SARIMA model was fitted. Subsequently, its residual series were inputted to the NNAR model. The nonlinear relationships that the residuals may contain can be mined adequately by neural networks. The final combined forecasting values of the time series were the sum of predictions from the SARIMA model and the adjusted residuals from NNAR model. The structure of the hybrid model is shown in Figure 1.

**Figure 1:**
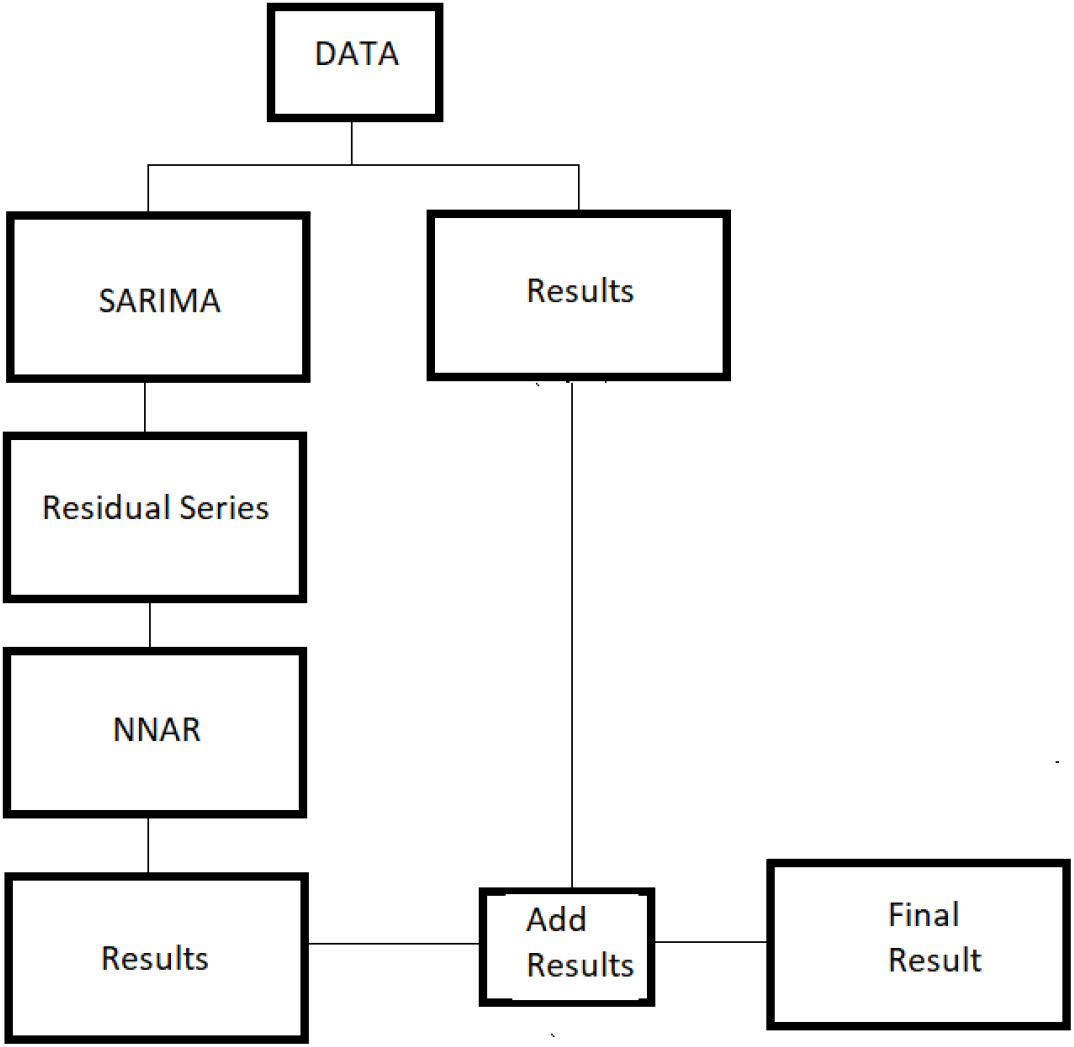
Structure of Hybrid Model

#### 2.2.4. Forecast evaluation methods

The performance of the model is related to the similarity in the forecast values for the test data and observed values. Three different forecast consistency measures are used for comparing the performances obtained for the SARIMA, NNAR and SARIMA-NNAR models: mean square error(MSE), mean absolute error(MAE) and mean absolute percentage error(MAPE). The smaller the MSE, MAE, and MAPE of the prediction model, the better its prediction accuracy. In the other words, the prediction model with the smallest MSE, Mae, and MAPE can be selected as the optimum models.

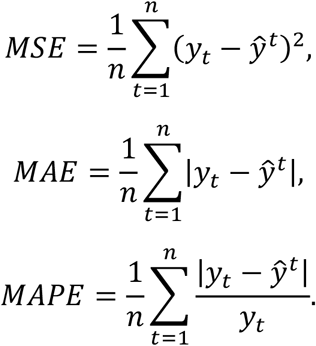

## 3. Results

In Turkey, 35608 people died due to the COVID-19, until April 17, 2021. Precautions are the most important means of reducing these deaths. If the death rates can be predicted forward, the governments can move faster in taking action. Covid-19 cases in Turkey increased in April 2020. The data used in the analysis are from March 11, 2020 to February 22, 2021.

However, the precautions taken have led to a decrease in the cases. When the precautions relaxed, it increased again. The time-series distribution of those who died from COVID-19 on a daily basis is given in Figure 2. This chart also includes ACF and PACF charts. According to these values, p and q values are estimated in the time series model. Since the ACF chart is showing a slow decline, it can speak of a trend in time series.

**Figure 2:**
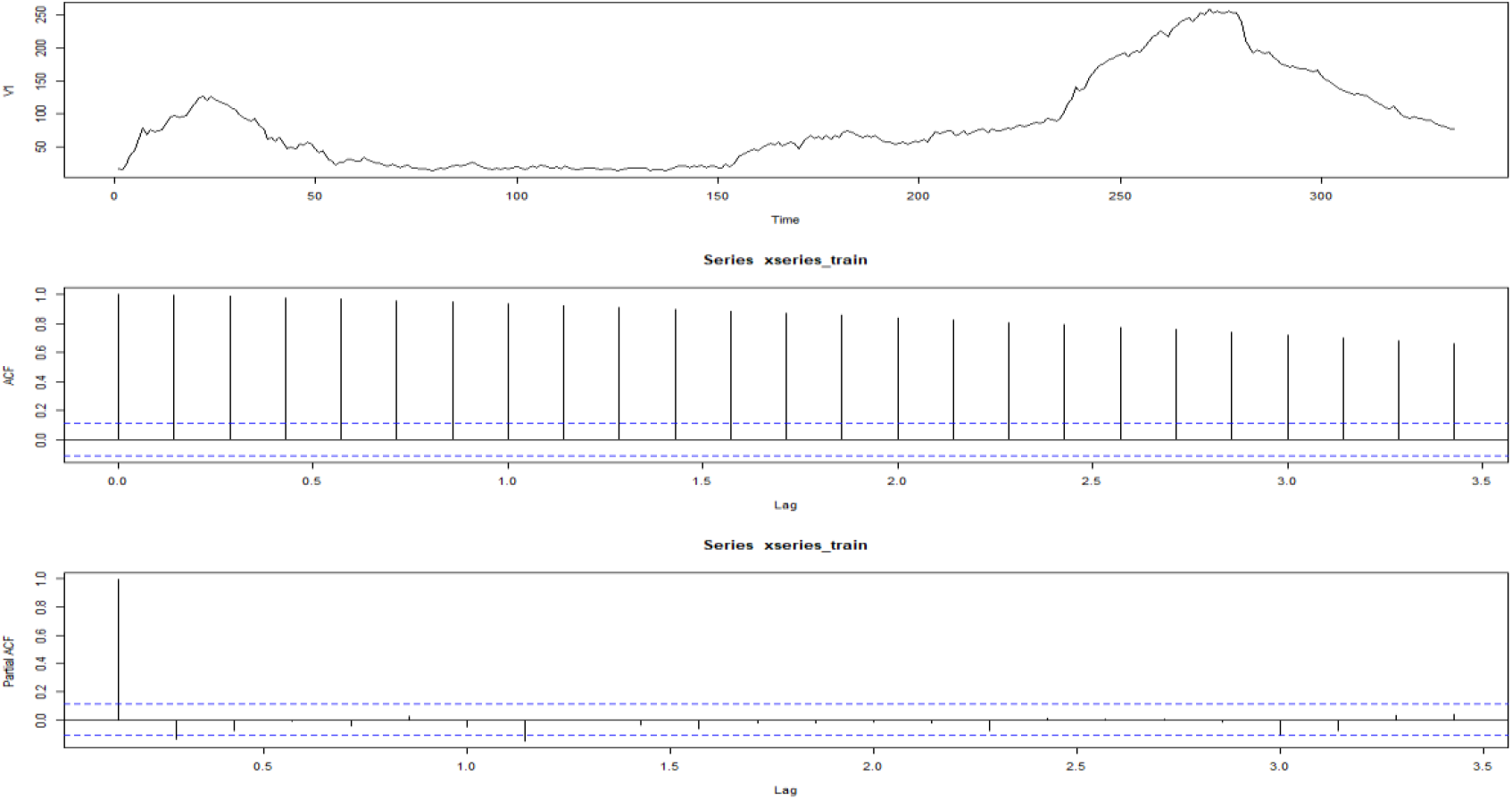
Time series and ACF-PACF plot

In Figure 3, the *decomposition()* function is divided into time series components to examine whether there is seasonality. The components of the series are given in figure 3.

**Figure 3:**
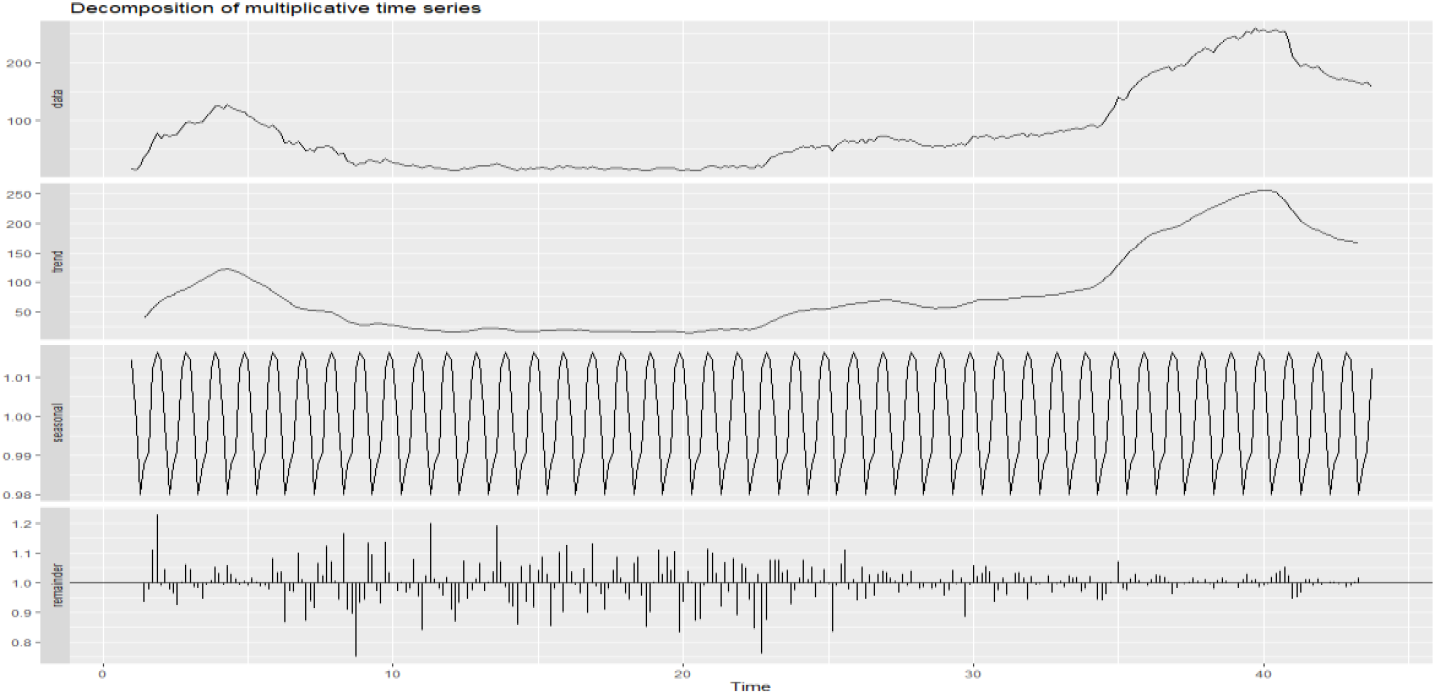

Time series analysis was first applied to the time series of those who died due to COVID-19. In the decomposition, only the trend is clearly separated, but we do not have any information for the p and q values. The ADF test states that the time series is not stationary (ADF=-13.737, p=0.01). For this reason, the most suitable model has been determined with the *auto*.*arima ()* function. In this case, SARIMA(2,1,2)(2,0,1)_7_ model was obtained as the most suitable model with *auto*.*arima*() function of forecast library [9]. The ACC_C_ value of the model is 1784.77 and the BIC value is 1813.88. The residuals of this model, ACF, and Normality analysis graph are given in Figure 4. Whether the residuals of the created model show autocorrelation was examined by Ljung-Box test and it was found that the residues did not show autocorrelation (χ^2^=14.476, df=24, p=0.9352).

**Figue 4:**
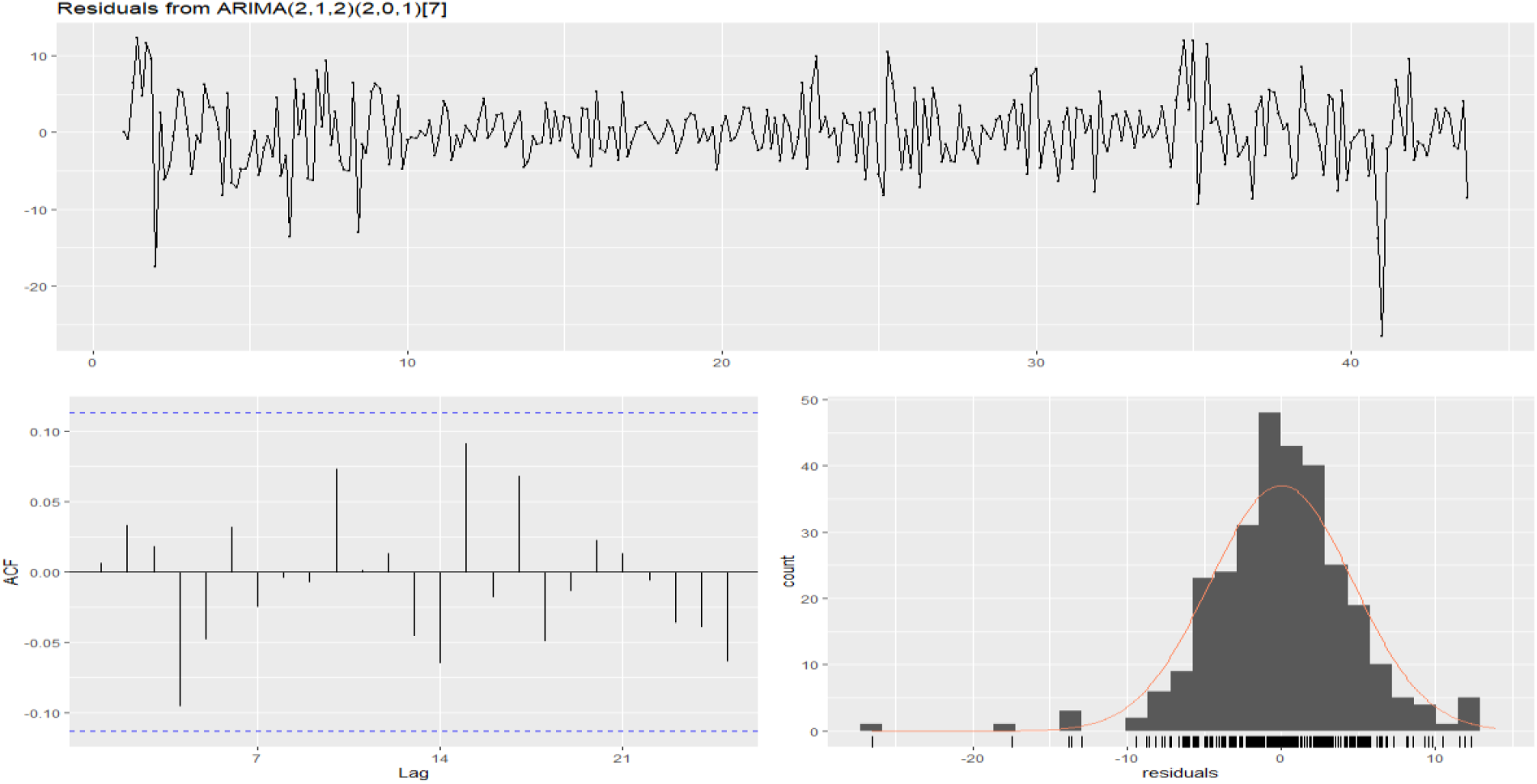
Residual plot, corresponding ACF plot and histogram from ARIMA(2,1,2)(2,0,1)_7_

### Best performing NNAR model

Due to seasonality, p = 1 was set. Again, the model of the forecast library was obtained automatically with the *nnetar* function. The model obtained by the function is the *nnar*(2,1,2)_7_ model. In addition, *nnar* models with different p and q values from 1 to 10 were obtained. Obtained models are listed in Table 1. Here, there are 9 models with the lowest RMSE value. Considering three performance indices in both training and validation sets, NNAR (10,1,7)_7_ was chosen as the most suitable model. Here, 70% of the data is divided into training data set and 30% as test data set. The graph of residuals, ACF, and Normality analysis of this model is given in Figure 5. Whether the residuals of the created model show autocorrelation were examined by Ljung-Box test and it was found that the residues did not show autocorrelation(χ^2^=14.775, df=24, p=0.9272).

**Table 1:**

|  | RMSE | MAE | MAPE | RMSE | MAE | MAPE |
| --- | --- | --- | --- | --- | --- | --- |
| (8,1,5)<sub>7</sub> | 3.723869 | 2.868839 | 5.960852 | 0.3500508 | 0.24699 | 0.24328 |
| (8,1,6)<sub>7</sub> | 3.612052 | 2.793796 | 5.86323 | 0.2381594 | 0.16410 | 0.15624 |
| (8,1,7)<sub>7</sub> | 3.559917 | 2.775637 | 5.850855 | 0.1829261 | 0.13529 | 0.13222 |
| (9,1,5)<sub>7</sub> | 3.602116 | 2.775739 | 5.853346 | 0.1111011 | 0.08418 | 0.08819 |
| (9,1,6)<sub>7</sub> | 3.486982 | 2.682023 | 5.732168 | 0.0888780 | 0.06401 | 0.06719 |
| (10,1,4)<sub>7</sub> | 3.705423 | 2.829739 | 5.903758 | 0.03699127 | 0.02864 | 0.03179 |
| (10,1,5)<sub>7</sub> | 3.495667 | 2.661528 | 5.67319 | 0.03167837 | 0.02561 | 0.02743 |
| (10,1,6)<sub>7</sub> | 3.383682 | 2.58536 | 5.569271 | 0.02092535 | 0.01722 | 0.01825 |
| (10,1,7)<sub>7</sub> | 3.2764 | 2.498271 | 5.460094 | 0.02042849 | 0.01772 | 0.01863 |

**Figue 5:**
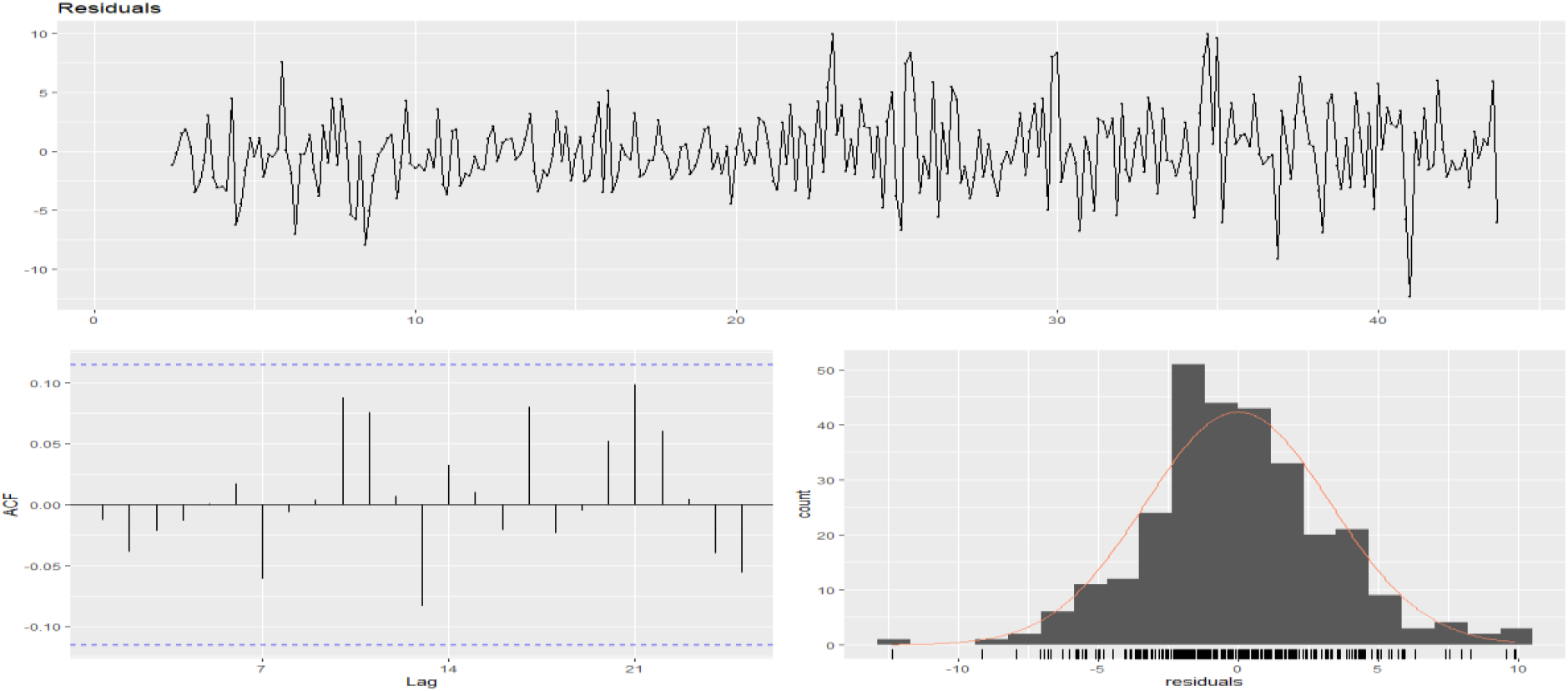
Residual plot, corresponding ACF plot and histogram from NNAR(10,1,7)_7_

### Best performing SARIMA - NNAR model

The SARIMA-NNAR model was obtained from the *forecastHybrid* library with the *hybridModel* function. Here, the SARIMA model is the model obtained by *auto*.*arima* function and the NNAR model is determined as NNAR (3,1,2)_7_. The graph of residuals, ACF, and Normality analysis of this model is given in Figure 6. Whether the residuals of the created model show autocorrelation were examined by Ljung-Box test and it was found that the residues did not show autocorrelation (χ^2^=10.602, df=24, p=0.9559).

**Figure 6:**
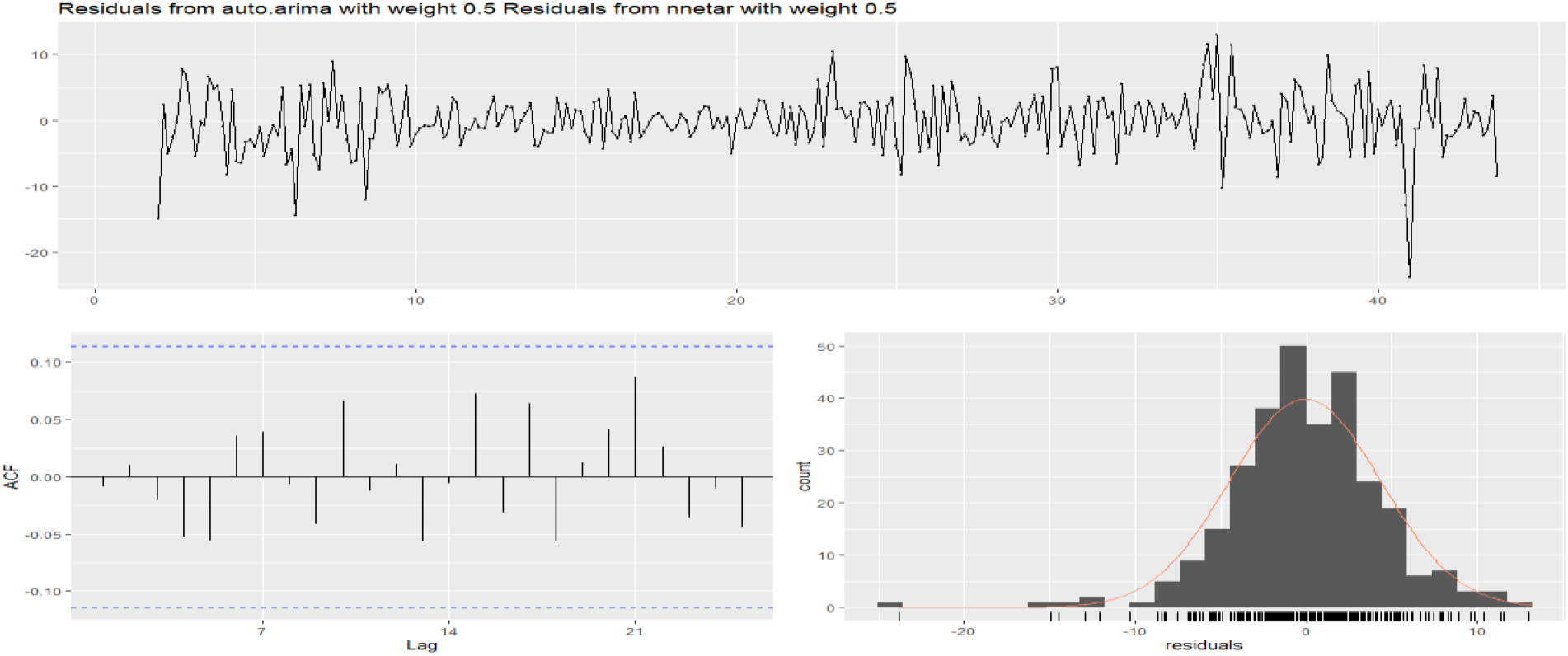
Residual plot, corresponding ACF plot and histogram from SARIMA-NNAR

The performances of the SARIMA, NNAR, and SARIMA-NNAR models tested in the study are given in Table 2. According to the table, the RMSE, MAE, and MAPE values of the NNAR model have the lowest values in the training set and the validation set.

**Tablo 2:**
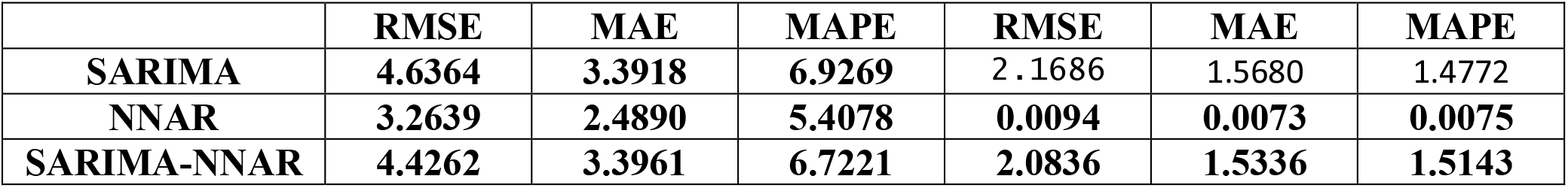
Accuracy of training set and 52 weeks forecasting in validation set.

**Table 3:** Twenty four days forecast

| Day | 1 | 2 | 3 | 4 | 5 | 6 | 7 | 8 | 9 | 10 | 11 | 12 |
| --- | --- | --- | --- | --- | --- | --- | --- | --- | --- | --- | --- | --- |
| Forecast | 74.75 | 73.22 | 69.94 | 68.91 | 67.47 | 66.21 | 65.11 | 63.64 | 62.25 | 61.05 | 60.10 | 59.36 |
| Day | 13 | 14 | 15 | 16 | 17 | 18 | 19 | 20 | 21 | 22 | 23 | 24 |
| Forecast | 58.53 | 57.77 | 56.99 | 56.23 | 55.63 | 55.04 | 54.46 | 53.89 | 53.30 | 52.73 | 52.15 | 51.57 |

The training and testing time series of the model of the three models obtained in the study is shown in Figure 7. It seems that all models fit well in terms of the training set. Since it would be misleading to look at it visually, the best model was obtained as the NNAR model according to the comparison criteria given in Table 1. The forecast for the next 24 days according to the NNAR model is given in Figure 8. Accordingly, it is seen that the number of deaths from COVID 19 will increase in the coming days.

**Figure 7:**
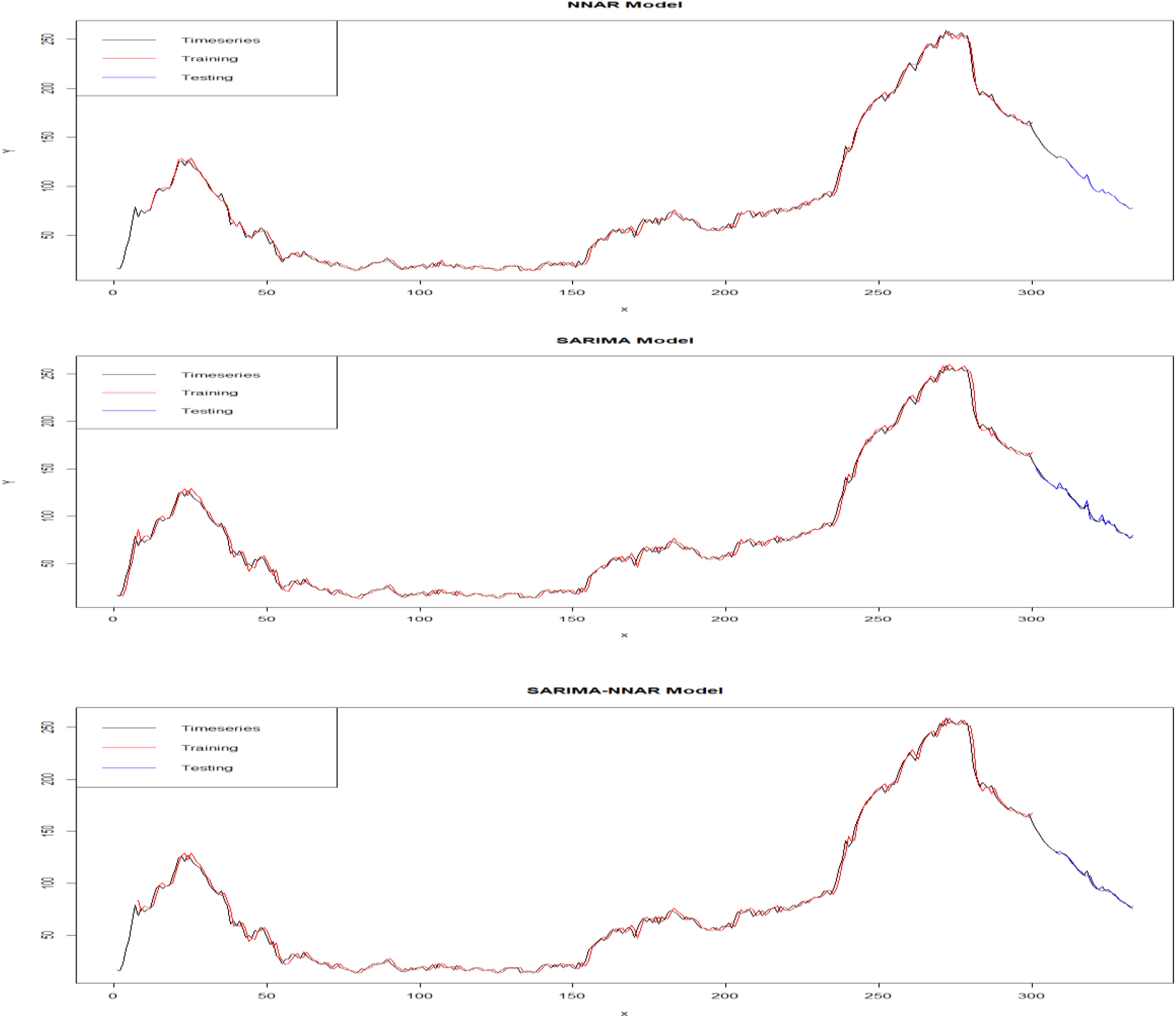
Fitted and 52 weeks forecasting time series plot of three models.

**Figure 8:**
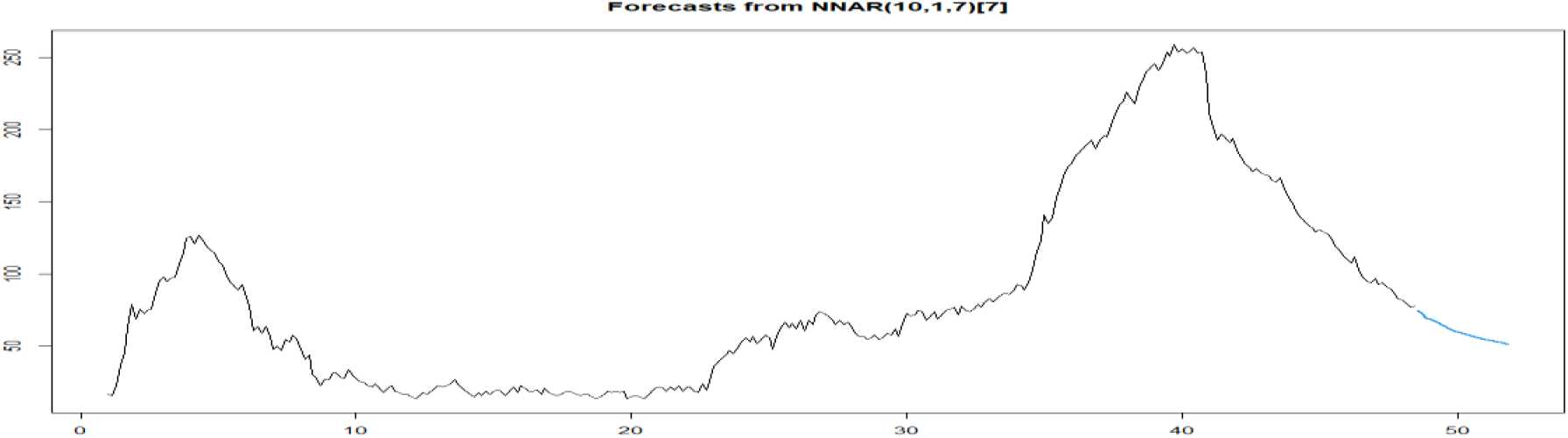
Prediction with NNAR (10,1,7) 7 model

## 4. Discussion

In this study, in Turkey, to estimate the number of deaths from COVID-19, SARIMA, NNAR, and SARIMA-NNAR models are used. It has been determined that the model that gives the best estimation result among these models and obtains the best result according to the comparison criteria of the models is NNAR. While developing the NNAR model, various simulations were made to try to predict the best model. In Figure 7, in the comparison of the three models, it was not observed that there was no serious difference between the models. However, the most suitable model for the result was determined according to the comparison criteria in Table 2. The second-best model was determined as the SARIMA-NNAR model. Using the NNAR model, it is possible to predict the number of deaths from COVID-19 in the future and take necessary measures accordingly. Thanks to these estimates and measures, the number of deaths can be reduced.

However, there are some limitations to this study. The first of these limitations is where the original data is taken. These data were obtained from the COVID-19 Information Page of the Republic of Turkey Health Ministry. This data can be included the possibility of false reporting and negligent reporting. The quality of the data can affect the build process and performance of the model to some extent. However, in order to obtain a better model, the best model was tried to be obtained by examining the p and q values from 1 to 10 instead of using an automatic model in the NNAR model. This model could also be achieved with SARIMA. In general, this process has not been tried, as the NNAR model gives better results than the SARIMA model.

## 5. Conclusions

A SARIMA, NNAR and SARIMA-NNAR models were developed to forecast COVID-19 disease cases in Turkey. In this study, from March 11, 2020, to February 22, 2021, in Turkey to estimate the number of cases of deaths from COVID-19 different models were tested and determined to be the most appropriate model NNAR model. This model is the model that gives the most appropriate result according to the comparison criteria. The NNAR model is the most suitable model for estimating the number of deaths and will give managers an idea to prevent and control the increase in deaths.

## Data Availability

Republic of Turkey Health Ministry, COVID-19 Information Page, https://covid19.saglik.gov.tr

## Data Availability Statement

All data generated or analyzed during this study are included in this published article. The dataset (the per day number of COVID-19 cases from March 11, 2020 to February 22, 2021) used and/or analyzed during the current study were obtained from web-site of Republic of Turkey of Health Ministry [7]

